# *Helicobacter pylori* infection reduces likelihood of death from advanced cancer: A longitudinal analysis from the Japanese DAIKO prospective cohort study

**DOI:** 10.1101/2021.12.02.21267212

**Authors:** Satoshi S. Nishizuka, Masahiro Nakatochi, Yuka Koizumi, Asahi Hishida, Rieko Okada, Sayo Kawai, Yoichi Sut­oh, Keisuke Koeda, Atsushi Shimizu, Mariko Naito, Kenji Wakai

## Abstract

**Background:** Paradoxically, patients with advanced stomach cancer who are *Helicobacter pylori*-positive (HP^+^) have a higher survival rate than those who are HP^-^. This finding suggests that HP infection has beneficial effects for cancer treatment. Present study examines whether HP^+^ individuals have a lower likelihood of death from cancer than those who are HP^-^.

**Methods and findings:** Prospective cohort data (*n* = 4,982 subjects enrolled in the DAIKO study between 2008-2010) was used to assess whether anti-HP antibody status as a surrogate for past-present HP infection was associated with cancer incidence. The median age in the primary registry was 53 years-old (range 34-69 years-old). Over the 8-year observation period there were 234 (4.7%) cancer cases in the cohort and 88 (1.8%) all-cause deaths. Urine anti-HP antibody data was available for all but one participant (n = 4,981; 99.97%). The number of HP^+^ and HP^-^ individuals was 1,826 (37%) and 3,156 (63%), respectively. Anti-HP antibody distribution per birth year revealed that earlier birth year was associated with higher HP^+^ rates. To remove confounding factors associated with birth year, a birth year-matched cohort (*n* = 3,376) was generated for subsequent analyses. All-cancer incidence was significantly higher in HP^+^ individuals than those who were HP^-^ (p=0.00328), whereas there was no significant difference in the cancer death rate between HP^+^ and HP^-^ individuals (p=0.888). Strikingly, we found that HP^+^ individuals who developed cancer had a better survival rate than would be expected based on cancer incidence. These results suggest that cancer patients who are HP^+^ may have a higher likelihood of survival than those who are HP^-^. Cox regression analysis for prognostic factors revealed that the hazards ratio of HP^+^ was 1.59-fold (95%CI 1.17-2.26) higher than HP^-^ in all-cancer incidence.

**Conclusions:** Potential systemic effects of HP^+^ status may contribute to reduced likelihood of death for patients with cancer.

**Data Availability Statement:** The data cannot be shared publicly as data sharing is not permitted according to Japanese Government data protection policies. Requests for data analysis may be accepted anonymously and conditionally upon IRB approval from Iwate Medical University and Nagoya University Graduate School of Medicine.

**Funding:** This study is supported by Grants-in-Aid for Scientific Research for Priority Areas of Cancer (No. 17015018); Grants-in-Aid for Innovative Areas (No. 221S0001); and the Japan Society for the Promotion of Science (JSPS) KAKENHI Grant (No. 19K09130 and No. 16H06277 [CoBiA]) from the Japanese Ministry of Education, Culture, Sports, Science and Technology (MEXT). The funders had no role in study design, data collection and analysis, decision to publish, or preparation of the manuscript.

**Competing interests:** The authors declare that no competing interests exist.

**Author summary:** *Why was this study done?:* > Although HP infection is a major cause of gastric diseases including cancer, how HP infection affects prolonged survival of advanced gastric cancer patients is unknown. > Reports of studies carried out in different countries and regions revealed that advanced gastric cancer patients who are HP^+^ exhibited prolonged post-treatment survival, even though the genetic background of patients, HP strains, and cancer treatment procedures differed. > Since most advanced gastric cancer patients underwent gastrectomy, the favorable prognosis of HP^+^ patients after multidisciplinary treatment may be due to putative systematic mechanisms associated with HP infection. > If putative systemic mechanisms associated with HP infection reduce the likelihood of death due to cancer, the cancer survival rate in the HP^+^ population should be lower than that for the HP^-^ population.

*What did the researchers do and find?:* > Using data from the DAIKO prospective cohort study in Nagoya, Japan, we analyzed the association between anti-HP antibody status, cumulative cancer incidence and all-cause and cancer-specific deaths. > The HP^+^ rate increased as birth year decreased. Thus, matching based on birth year between 1935 and 1975 was performed to correct for confounding factors associated with birth year. > Despite a significantly higher all-cancer incidence for HP^+^ individuals compared to those who were HP^-^, no difference in the all-cause and cancer death rate was observed between HP^+^ and HP^-^ individuals.

*What do these findings mean?:* > HP^+^ individuals are less susceptible to death relative to their incidence of cancer. > Patients with advanced stage cancer who are HP^+^ may have a better treatment response/tolerance than those who are HP^-^. > Additional longitudinal analyses are warranted to evaluate the effect of HP^+^ status on prolonged survival of patients with advanced-stage cancer.

## Introduction

*Helicobacter pylori* (HP) infection is considered to be an etiologic carcinogen of the stomach [1]. Paradoxically, however, investigators have recognized that the survival rate of patients with advanced gastric cancer who are HP^+^ is higher than that for HP^-^ patients [2–8]. Regarding HP infection in the context of post-operative adjuvant chemotherapy for patients with Stage II/III gastric cancer [9, 10], our previous study demonstrated that those patients who were HP^+^ had a clearly higher survival rate over the 10-year observation period [11]. In particular, patients who were HP^+^ and received adjuvant chemotherapy with S-1 showed >20% better overall survival than did those who were HP^-^, which was further confirmed by propensity score matching analysis. Stratified Cox proportional hazards regression analysis revealed that all 10 subgroups had better survival in the respective HP^+^ groups [11].

The beneficial effect of HP infection for advanced gastric cancer has been reported in studies carried out in a variety of countries that have varying HP genotype and diet as well as different therapeutic strategies for advanced gastric cancer [2–8, 11]. This result suggests that putative effects of HP infection may contribute to fundamental functions that prolong survival. Some systematic effects, most likely sustained immune system activation, could thus play a specific role in post-operative chemotherapy. Alternatively, HP infection could trigger non-specific enhancement of innate immune system activity to create a *de facto* heterologous immunological memory that has been termed trained immunity [12]. Such triggered immunity that has some anticancer effects has been shown in patients with high-risk non-muscular invasive bladder cancer for which intravesical instillation of Bacillus Calmette-Guérin (BCG) has been the gold-standard treatment [13]. For gastric cancer, immunotherapy using OK-432, a freeze-dried product prepared by incubating the low-virulence Su strain of group A *Streptococcus pyogenes* with penicillin [14], has been actively investigated [15]. Although not the gold-standard for gastric cancer, therapies like OK-432 that promote immune responses appear to suppress relapse in both humans and mouse models of gastric cancer [16–19]. If immune activation indeed plays a role in suppressing either the development or progression of cancer, then sustained HP infection should also show some beneficial effect in terms of cancer incidence and all-cause or cancer death in a longitudinal cohort.

The subject patients of most previous reports, including ours, had undergone gastrectomy, meaning that HP could no longer colonize the stomach. Thus, the observed effect of HP on prolonged survival could be systemic and perhaps lifelong, and thus may have an antitumor potential. Using data from the DAIKO prospective cohort in Nagoya, Japan, the present study aimed to understand whether the putative effect associated with HP infection has an antitumor effect with respect to reduction of incidence of all-cancer as well as breast, colorectal, lung, prostate, and stomach cancers. The effects of HP on all-cancer deaths were also assessed. Although the observation period of the DAIKO cohort to date is eight years, the beneficial effect of HP infection can be determined if the all-cancer death rate in HP^+^ individuals is lower than the all-cancer incidence in those who are HP^+^. This putative effect suggests that HP^+^ cancer patients have a lower likelihood of death compared to HP^-^ cancer patients.

## Methods

### Ethics statement

Ethical approval for the parent Japan Multi-Institutional Collaborative Cohort Study (J-MICC) study was granted by Nagoya University Graduate School of Medicine Institutional Review Board [20]. Based on the IRB approval by the Nagoya University Graduate School of Medicine, a separate ethical approval for the current analysis using the DAIKO study was granted by both Nagoya University Graduate School of Medicine IRB (2008-0618-2) and Iwate Medical University School of Medicine IRB (MH2018-019). The study was conducted according to Helsinki Declaration principles.

### Source cohort

The DAIKO study includes 5,165 participants aged 35-69 years-old who were residents of Nagoya and were enrolled between June 2008 and May 2010 at the Daiko Medical Center of Nagoya University, Nagoya, Japan. The J-MICC Study is the parental cohort of the DAIKO study, which includes 13 institutes across Japan with enrollment of >100,000. Follow-up surveys have been conducted since 2005 [20]; and the end of follow-up of the current study was Dec 16, 2016. The main objective of the J-MICC study is to investigate gene-environmental interactions with lifestyle-related diseases, mainly cancer. The cohorts have also provided opportunities to perform cross-sectional studies on lifestyle factors, biomarkers, and genotypes [21–25]. In the present study, we analyzed DAIKO study data in terms of cancer incidence and survival to verify whether these events were associated with anti-HP antibody status that indicates the immune status of an individual with respect to past/present HP infection.

### Study population

Participants in the present study were a subset of the DAIKO study, from whom informed consent was obtained upon enrollment in studies outside of the J-MICC-assigned program. Baseline data were obtained from 5,165 participants, and 183 participants were excluded for the following reasons: lack of consent to be a part of research studies except for the parent J-MICC study (n=170); ineligibility (n=12); and revocation of informed consent at baseline (n=1). Overall, 4,981 (96.4% of the J-MICC study participants) individuals were enrolled in the present study.

### Baseline variables

Baseline covariates of the present study included age, birth year, gender, body mass index, blood pressure, laboratory blood data, history of cancer, history of medication for HP, and urine anti-HP antibody status determined using an immunochromatography kit (RAPIRAN, Otsuka Pharmaceutical, Tokushima, Japan) [26–28].

### Outcome measures

The primary end point was all-cancer incidence, and all-cause death. The all-cancer incidence was further categorized into cancer types when appropriate. Analysis of individual cancer types included those of asynchronous secondary cancer. The classification of cancer types was defined using the International Statistical Classification of Diseases for Oncology, Third Edition (ICD-O-III). The vital and residential status of participants was determined by using the population registry. For logistical reasons, we ended the follow-up of subjects who had moved out of the study area (Nagoya city). Causes of death were confirmed based on the vital statistics data provided by the Ministry of Health, Labor and Welfare, Japan. Cancer incidence data was obtained from the Aichi Cancer Registry.

### Statistical analysis

In principle, all variables were compared based on the anti-HP antibody status defined in a binary manner (i.e., presence of antibody, HP-positive, HP^+^; absence of antibody, HP-negative, HP^-^). Categorical and continuous variables were analyzed using Fisher’s exact test and Wilcoxon’s rank sum test, respectively. Subject matching was performed to correct for birth year. To minimize the effects of a lack of background uniformity when comparing cancer incidence/death and anti-HP antibody status, the following procedure was performed when necessary: (a) Exclusion of participants who had a case history of any type of cancer; and (b) frequency matching according to birth year by gender. The proportion of event-free survival was estimated using the Kaplan-Meier method. A log-rank test was used to examine the null hypothesis whereby there was no difference in the proportion of event-free survival between the populations in terms of the probability of an event at any time point. A Cox proportional hazards model was used to estimate the hazards ratio (HR). The follow-up period was computed from the baseline survey to cancer incidence (in the analysis for cancer incidence), death of any cause, moving out of the study area, and the end of follow-up (Dec 31, 2016). The model included age, sex (1: male, 2: female), smoking status (1: current or ever, 0: never), and drinking habits (1: current or ever, 0: never). P-value <0.05 was considered statistically significant. All statistical analyses were performed using R.

## Results

### Participant characteristics

The population of the prospective DAIKO cohort study that was eligible for the present study (n=4,982) included 1,417 men and 3,567 women, with a median age of 53. Results of urine anti-HP antibody tests were available for 4,981 out of 4,982 (99.9%) participants. There were 1,825 (37%) HP^+^ individuals and 3,156 (63%) HP^-^ individuals. The summary of participant characteristics is shown in Table 1. Individuals who were HP^+^ tended to be male and older relative to HP^-^ individuals. HP^+^ status was also associated with higher BMI and history of smoking. Although the absolute difference was small, all clinical and laboratory continuous variables tested exhibited higher risk factors for major chronic diseases for HP^+^ compared to HP^-^ individuals (Table S1) [29, 30].

**Table 1.** Characteristics of study participants

|  |  | HP<sup>+</sup> (n=1,825) |  | HP<sup>-</sup> (n=3,156) |  |  |
| --- | --- | --- | --- | --- | --- | --- |
| Variable | Value | n | Freq | n | Freq | P value |
| Sex | Male | 652 | 0.308 | 854 | 0.271 | 0.00506 |
|  | Female | 1,263 | 0.692 | 2,302 | 0.729 |  |
| Age | Median | 58.6 (49.3, 64.7) |  | 50.3 (42.4, 60.5) |  | 3.47 x 10<sup>-60</sup> |
| BMI<sup>a</sup> | Median | 21.5 (19.7, 23.8) |  | 21.1 (19.4, 23.3) |  | 0.0000874 |
| Smoking | Yes | 609 | 0.334 | 962 | 0.305 | 0.034 |
|  | No | 1,215 | 0.666 | 2,194 | 0.695 |  |
| Drinking | Yes | 1,009 | 0.553 | 1,795 | 0.569 | 0.273 |
|  | No | 816 | 0.447 | 1,360 | 0.431 |  |

HP, *Helicobacter pylori*; BMI, Body mass index; Yes, current or ever;
No, never; <sup>a</sup>HP<sup>-</sup> (n=3,155).

### HP eradication

Nearly 90% of participants had never been assessed for HP prior to enrolling in the DAIKO study. Among the 472 (9.5%) participants who had undergone an HP examination (method unknown), 314 (66.5%) were positive, and 133 (28.2%) were negative. The status of 25 individuals (5.3%) was unknown or no credible information available. Among the previously-diagnosed HP^+^ participants (n=314), 152 (48.4%) were anti-HP antibody positive (HP^+^) while 162 (51.6%) were HP^-^ at the DAIKO baseline. Among the previously-diagnosed HP^-^ participants (n=133), 26 (19.5%) were HP^+^ while 107 (80.5%) were HP^-^ at DAIKO baseline. Thus, the consistency rate was 57.7% among any previous HP exam and the present urine anti-HP antibody test. It should be noted that HP^-^ participants at the DAIKO baseline could include those for whom HP was successfully eradicated. In fact, among the previously-HP^+^ participants, 162 (51.6%) had successful eradication. The self-claimed “successful eradication” resulted in 53 (33.1%) anti-HP antibody positive subjects, whereas 109 (66.9%) were negative at baseline. No definitive time point for eradication was available for the present cohort.

### Birth year effect

The number of HP^+^ individuals categorized by birth year was 76 (1.5% of all HP^+^ cases) for 1938-1939, 876 (17.6%) for 1940-1944, 793 (15.9%) for 1945-1949, 617 (12.4%) for 1950-1954, 645 (12.9%) for 1955-1959, 706 (14.2%) for 1960-1964, 629 (12.6%) for 1965-1969, 630 (12.6%) for 1970-1974, and 9 (0.18%) for 1975. There was a clear trend that lower birth year was associated with higher rate of HP^+^ (Fig. 1). The mean HP^+^ fraction by birth year 1938-1944 was 0.58, whereas that for 1970-1975 was 0.21. The fraction of females with HP^+^ and birth year 1938-1944 was 0.45, whereas that for 1970-1975 was 0.15. The HP^+^ fraction for men was higher than that for women across the birth years. The birth year effect by sex is shown in Figure 1.

**Fig 1.**
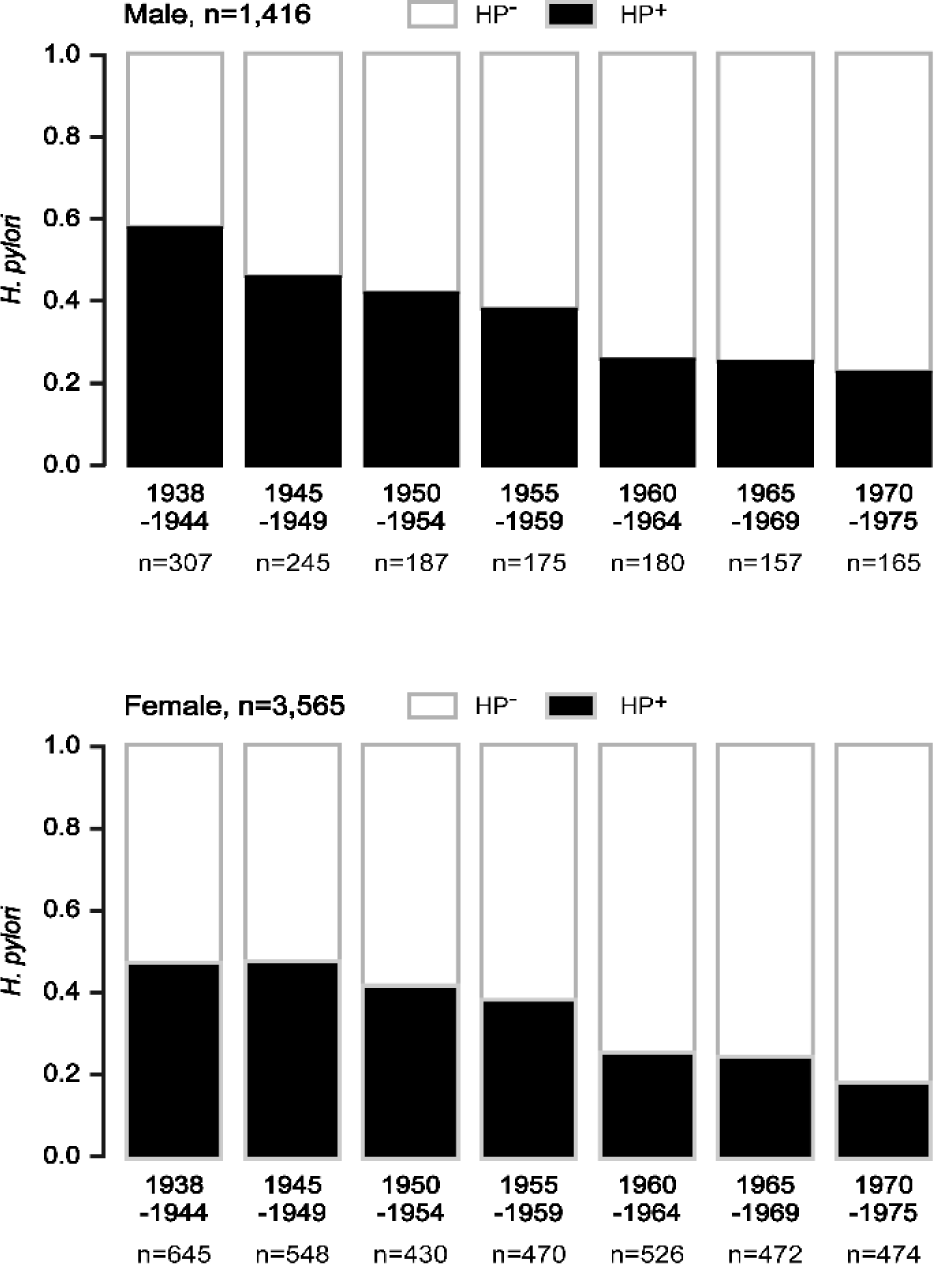
Anti-HP antibody status by birth year. The HP^+^ rate is shown with five-year bins according to birth year. Black areas in each bar represent the HP^+^ fraction.

### Matching

The HP^+^ participants were frequency-matched to HP^-^ respondents for five-year bins of birth year and gender to minimize confounding factors. Those subjects who had a previous history of cancer were excluded. With birth year matching, the categorical patient characteristics showed no significant difference between HP^+^ and HP^-^ individuals (Table 2). Moreover, after matching, almost all continuous variables showed no difference between HP^+^ and HP^-^ individuals except for the HDL cholesterol level for which the estimated mean difference was 3 mg/dl (Table S2).

**Table 2.** Characteristics of participants after matching

|  |  | HP<sup>+</sup> (n=1,688) |  | HP<sup>-</sup> (n=1,688) |  |  |
| --- | --- | --- | --- | --- | --- | --- |
| Variable | Value | n | Freq | n | Freq | P value |
| Sex | Male | 489 | 0.290 | 489 | 0.290 | 1.00 |
|  | Female | 1,199 | 0.710 | 1,199 | 0.710 |  |
| Age (yrs) | Median | 57.9 (48.6, 64.1) |  | 57.8 (48.8, 63.9) |  | 0.866 |
| BMI<sup>a</sup> | Median | 21.5 (19.7, 23.8) |  | 21.4 (19.7, 23.5) |  | 0.37 |
| Smoking | Yes | 545 | 0.323 | 522 | 0.309 | 0.395 |
|  | No | 1,142 | 0.677 | 1,166 | 0.691 |  |
| Drinking | Yes | 920 | 0.545 | 968 | 0.573 | 0.103 |
|  | No | 768 | 0.455 | 720 | 0.427 |  |

HP, *Helicobacter pylori*; BMI, Body mass index; Yes, current or ever; No, never; <sup>a</sup>HP<sup>-</sup> (n=1,687)

### Cancer incidence

The cancer incidence was 105 and 67 in HP^+^ (n=1688) and HP^-^ (n=1688) individuals, respectively (p=0.00367, Fisher’s exact test). The individual cancer types that had significantly higher incidence in the HP^+^ population were both gastric (20 and 5 in HP^+^ and HP^-^ individuals, respectively) and non-gastric (87 and 62 in HP^+^ and HP^-^ individuals, respectively). Other cancer types including uterine, lung, prostate, colon, and breast showed no significant difference in cancer incidence in terms of HP infection status, although the number of cases was too small to reach a definitive conclusion about the effect of HP status. The all-cause death incidence in terms of HP status also showed no significant difference. The cancer incidence after matching is shown in Table S3.

### Survival analysis

The crude cumulative all-cancer incidence rate was higher for HP^+^ individuals than that for HP^-^ (n=3,376, P=0.0038; Fig. 2). As expected, the incidence of gastric cancer was significantly higher in HP^+^ individuals than those who were HP^-^ (p=0.0027; Fig. 3A). Interestingly, however, the incidence of non-gastric cancer was also significantly higher in HP^+^ individuals than those who were HP^-^ (p=0.039; Fig. 3B). According to cancer type, the cumulative cancer incidence was also assessed for lung, colorectal, rectum, prostate, and breast cancer (S1-5 Figs).

**Fig 2.**
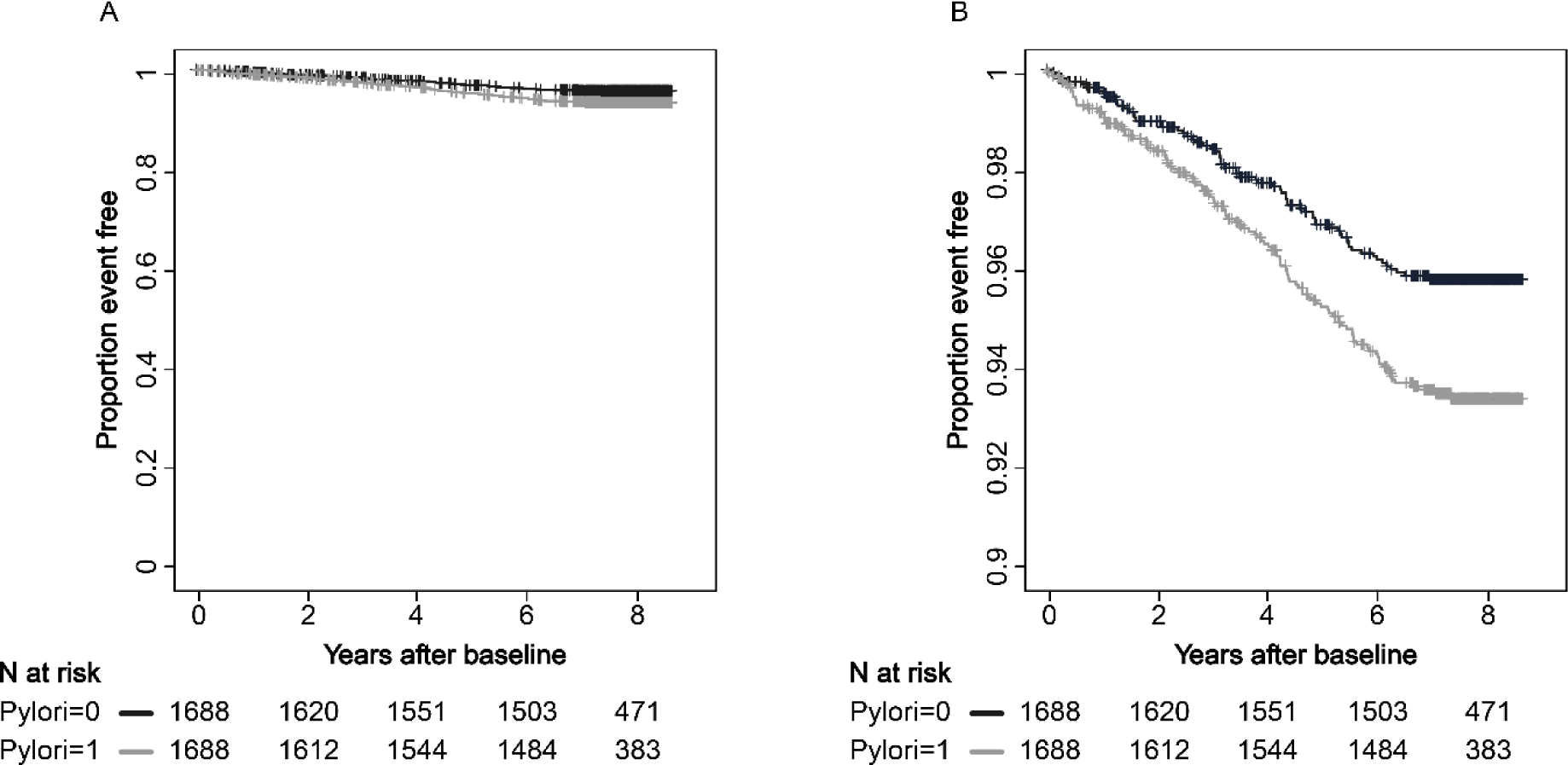
All cancer incidence. Kaplan-Meier estimation of all cancer incidence stratified by anti-HP antibody status. A, axis range 0.00-1.00; B, axis range 0.90-1.00.

**Fig 3.**
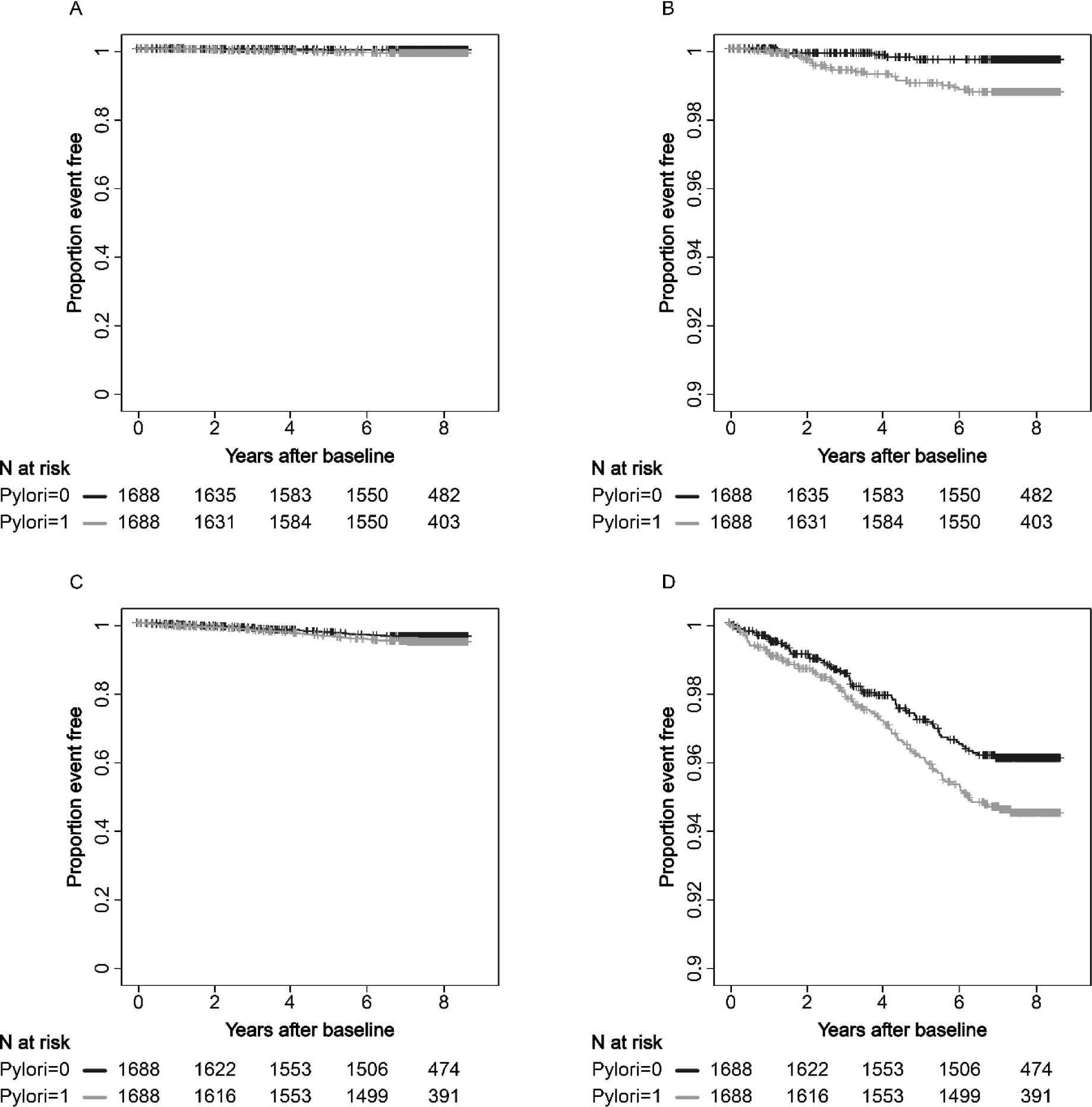
Gastric and non-gastric cancer incidence. Kaplan-Meier estimation of gastric (A, B) and non-gastric (C, D) cancer incidence stratified by anti-HP antibody status. A and C, axis range 0.00-1.00; B and D, axis range 0.90-1.00.

The cumulative all-cause death rate was not significantly different between HP^+^ and HP^-^ individuals (p=0.972; Fig. 4A). Although a limited number of deaths were recorded (n=33 out of 3376 matching cases), cancer-specific deaths did not differ between HP^+^ and HP^-^ individuals (n=3376, p=0.888; Fig. 4B).

**Fig 4.**
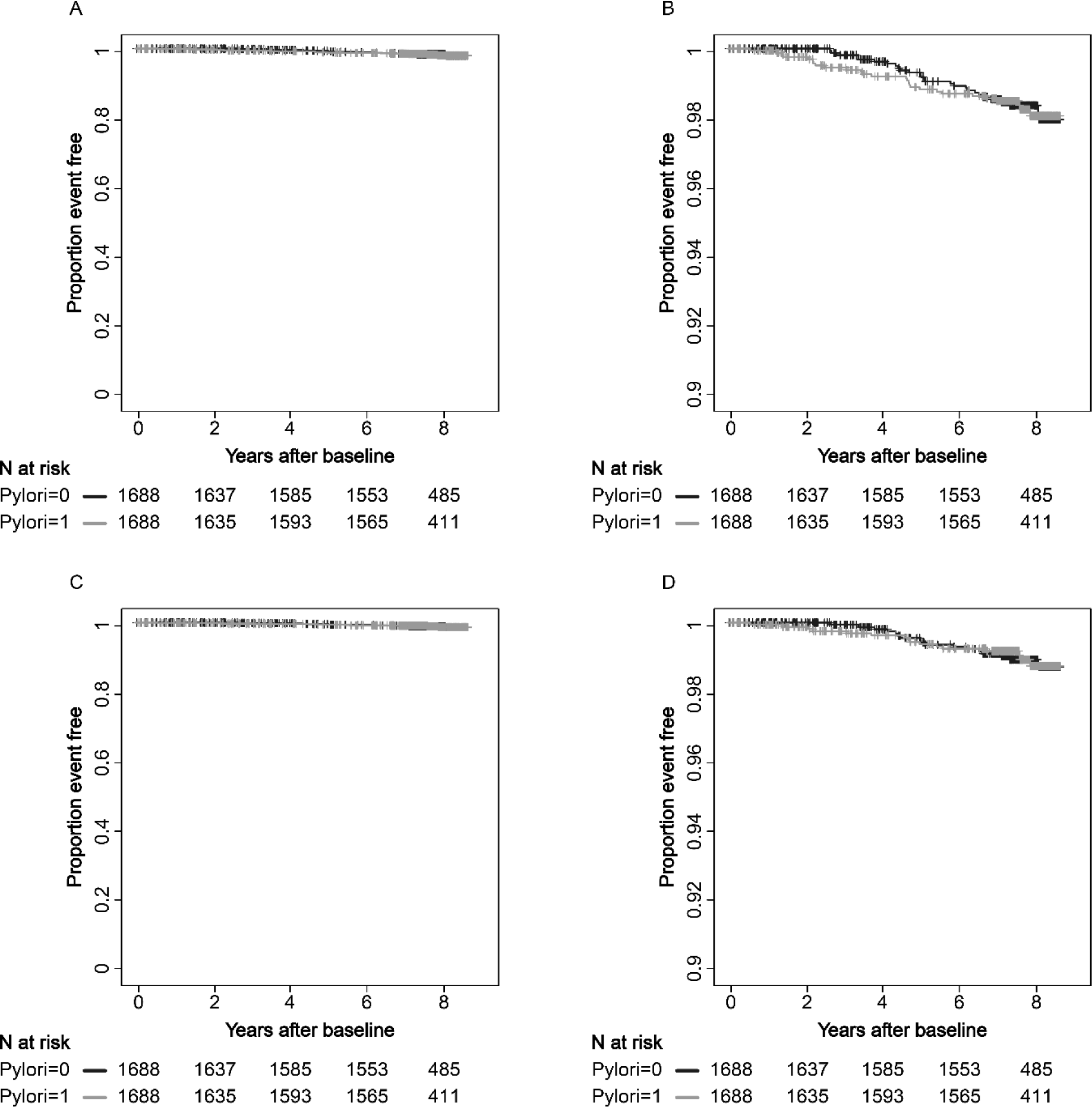
All-cause and cancer-specific deaths. Kaplan-Meier estimation of all-cause deaths (A, B) and deaths due to cancer (C, D) stratified by anti-HP antibody status. A and C, axis range 0.00-1.00; B and D, axis range 0.90-1.00.

### Predictor variables

To identify independent predictive variables for the risk of incidence for all-cancer, gastric, and non-gastric cancer, a Cox proportional-hazards model was applied with variables including age, sex, drinking, smoking, and HP infection. The hazards ratio of all-cancer incidence by HP was 1.59 (p=0.00297), which was lower than the hazard ratio for smoking (1.97; p=0.000422) but higher than that for alcohol consumption (1.24; p=0.216, Table S3). The hazards ratio for the gastric cancer incidence was 3.93 (p=0.00631, Table S4), whereas that for non-gastric cancer incidence in HP^+^ over HP^-^ was 1.42 (p=0.0356, Table S5). The number of gastric cancer cases was 20 and 5 in HP^+^ (n=1,688) and HP^-^ (n=1,688) individuals, respectively, which was too small to allow additional modeling with respect to other cancer types in the current study observation period.

## Discussion

In our study cohort from the DAIKO study, we demonstrate that the cancer incidence of the HP^+^ population was significantly higher than the HP^-^ population. However, the all-cancer death rate for the HP^+^ population was similar to that for the HP^-^ population. Two possible hypotheses can be drawn from these observations. First, the HP^+^ population appears to be more susceptible to cancer. Second, HP^+^ populations may have higher response or sensitivity to treatment for advanced cancer. Important premises can explain these hypotheses including establishment of HP^+^ infection early in life [31–34] and therapy for advanced cancer in Japan is almost always attempted before death as part of coverage by the Japanese universal health insurance system, *kaihoken* [35]. Therefore, the present results suggest that HP infection may have triggered acquisition of innate immunity that is beneficial for advanced cancer therapy [12].

With respect to the carcinogenic potential of HP, it has been suggested that HP can be largely divided into Western and East Asian strains based on genetic polymorphisms of EPIYA motif(s) encoded by the SHP-2 domain of the *cagA* gene [36, 37]. The EPIYA motif(s) of the CagA protein is phosphorylated and in Eastern Asian strains CagA has greater involvement in SHP-2 binding *in vitro* compared to Western strains [38]. Polymorphisms of EPIYA motif(s) of the CagA protein are known to be associated with a high incidence of gastric cancer [36, 39]. Moreover, across the last 20 years, evidence obtained in a series of large-scale clinical studies on peri- and/or post-operative therapy has established diverse therapeutic protocols that vary across regions worldwide [9, 10, 40, 41]. These geographical regions are largely linked to ethnic groups, and ethnic group interacts with virulence genotypes of HP [42]. From the perspective of the host, Toll-like receptors respond to HP infection and trigger the innate adaptive immune response [43]. Moreover, the gene expression profile of post-HP eradication gastric mucosa is altered in a variety of functions [44]. Cancer development can occur over several decades after HP infection and a fully-controlled study throughout the lifetime of an individual can be challenging [45, 46].

Epidemiological approaches may at least compensate to support the postulated mechanisms. Therefore, the favorable survival reported for patients with advanced cancer who are HP^+^ in studies carried out in different geographic regions suggests that the presence of HP infection may predominate over the diversity of factors such as treatment approach, ethnicity, and HP strain, and is probably supported by “trained innate immunity” [12].

After recognition of a pathogen, one of the ultimate purposes of disease control is to reduce deaths caused by the pathogen. For gastric cancer, eradication of HP has been considered to reduce a variety of gastric diseases including gastritis, peptic ulcer, and potentially gastric cancer [47–49]. However, the role of HP in gastric carcinogenesis may be limited to the development of “early” gastric cancer, in which normal mucosal cells transform to a malignant phenotype [50, 51]. More advanced-stage gastric cancer cases have substantial invasion beyond the submucosae where cancer cell division is no longer regulated by the malignant signal transduction induced by HP, which has been well presented in the “hit-and-run” model [52].

In Japan, the 5-year disease-free survival of pT1N0M0 gastric cancer is 99%, and that for pT1N1M0/pT2N0M0 is 94% and 92%, respectively [53]. Thus, early gastric cancer may not be prioritized in terms of efforts to reduce gastric cancer-deaths. With the universal health policy in Japan [35], nearly all advanced cancer patients can be assumed to have been treated at least once in a hospital. It is clear that nearly all gastric cancer-deaths are caused by advanced stage or recurrent gastric cancer. Therefore, the beneficial effect of HP in these gastric cancer patients should be considered differently from the simple malignant transformation of gastric mucosal cells. Here we suggest that stratification of treatment for these advanced, including non-gastric, cancer patients by HP infection status in which promoting therapy or maintaining alternative regimens for the HP^-^ population might be a prompt practical option.

The prevalence of HP in some regions of the world remains high [54]. The eradication of HP appears to be associated with a reduced incidence of gastric cancer [55]. It is, however, important to consider the fact that gastric cancer death rate has been continuously decreasing in many ethnic groups and countries including Japan since 1970s, without a concerted nationwide eradication program [56–60]. In the present study, we demonstrated that anti-HP antibody status was associated with birth year in that those having later birth year had a lower HP^+^ rate. These findings suggest that the role of HP in cancer treatment is one of the treatment stratification indices for advanced gastric cancer patients in most countries, whereas the number of gastric cancer deaths will likely continue to decrease naturally without efforts for HP eradication.

The present study has some limitations. The observational period is eight years, and the mean age at baseline was 58. A longer observation period would increase the cancer incidence and number of deaths. In fact, the number of all-cancer deaths may be too small to be conclusively analyzed as an independent parameter. We would have more evaluable cases in terms of tumor-type specific HP effect other than gastric cancer. The HP eradication history was reported by individual participants as personal medical records were not fully accessible for the present cohort. The HP infection was evaluated by only testing for anti-HP antibody in the urine. The anti-HP antibody status may be considered to reflect past or present HP infection, but no information about whether urine antibody levels had naturally attenuated was available. Therefore, this parameter might be less robust than extracting positive results from multiple HP examinations. The HP strain was not assessed, although most HP strains in Japan are CagA^+^ [61]. Depending on the practicality and objectives, detailed characterization of HP may be warranted in future studies.

## Conclusions

HP infection is associated with susceptibility to diverse types of cancer. However, HP infection has a favorable survival effect for patients with advanced cancer. With previous reports showing a better prognosis of advanced gastric cancer patients who are HP^+^ than those for HP^-^ patients from various regions of the world [2–8, 11] where the therapeutic regimens are diverse, HP infection may be a predominant factor for survival of not only patients with gastric cancer, but also for those with other types of cancer.

## Data Availability

The data cannot be shared publicly as data sharing is not permitted according to Japanese Government data protection policies. Requests for data analysis may be accepted anonymously and conditionally upon IRB approval from Iwate Medical University and Nagoya University Graduate School of Medicine.

## Acknowledgements

We thank all the DAIKO study organizers, staff, and participants.

## Author Contributions

**Conceptualization:** Satoshi Nishizuka.

**Data curation:** Masahiro Nakatochi, Asahi Hishida, Rieko Okada, Sayo Kawai.

**Formal analysis**: Masahiro Nakatochi, Kenji Wakai.

**Funding acquisition**: Keisuke Koeda, Satoshi Nishizuka, Kenji Wakai.

**Investigation**: Satoshi Nishizuka, Masahiro Nakatochi. **Methodology**: Satoshi Nishizuka, Masahiro Nakatochi, Yoichi Sutoh, Atsushi Shimizu, Mariko Naito, Kenji Wakai.

**Supervision**: Satoshi Nishizuka, Masahiro Nakatochi, Mariko Naito, Kenji Wakai.

**Visualization**: Masahiro Nakatochi, Yuka Koizumi, Kenji Wakai.

**Writing – original draft**: Satoshi Nishizuka.

**Writing – review & editing**: Satoshi Nishizuka, Masahiro Nakatochi, Mariko Naito, Kenji Wakai.

## Abbreviations

HP: Helicobacter pylori
CI: confidence interval
J-: MICC Japan Multi-Institutional Collaborative Cohort Study.

## Supporting information

**S1 Table.** Continuous variables for the participants

| Variable (unit) | HP<sup>+</sup> (n=1,825) | HP<sup>-</sup> (n=3,156) | P value |
| --- | --- | --- | --- |
| Age (yr) | 58.6 (49.3, 64.7) | 50.3 (42.4, 60.5) | 3.47 x 10<sup>-60</sup> |
| Waist (cm)<sup>a</sup> | 80.8 (74.8, 87) | 78.6 (72.8, 85) | 6.04 x 10<sup>-12</sup> |
| SBP (mmHg) | 117.5 (105.5, 133) | 113.5 (103, 127.5) | 3.24 x 10<sup>-12</sup> |
| DBP (mmHg) | 71.5 (64, 81.5) | 70.5 (63, 79.5) | 0.000289 |
| TC (mg/dL)<sup>b</sup> | 210 (188, 235) | 205 (182, 229) | 5.91 x 10<sup>-8</sup> |
| TG (mg/dL)<sup>b</sup> | 83 (57.8, 119) | 74 (54, 109) | 5.87 x 10<sup>-8</sup> |
| HDL (mg/dL)<sup>b</sup> | 61 (52, 73) | 65 (55, 76) | 2.60 x 10<sup>-10</sup> |
| AST (GOT) (IU/L)<sup>b</sup> | 20 (17, 24) | 19 (17, 23) | 2.68 x 10<sup>-6</sup> |
| ALT (GPT) (IU/L)<sup>b</sup> | 16 (12, 21) | 15 (11, 20) | 0.00893 |
| γ-GTP (IU/L)<sup>b</sup> | 20 (14, 32) | 18 (13, 29) | 6.15 x 10<sup>-6</sup> |
| Cr (mg/dL)<sup>b</sup> | 0.6 (0.6, 0.7) | 0.6 (0.6, 0.7) | 0.00703 |
| UA (mg/dL)<sup>b</sup> | 4.7 (4, 5.7) | 4.5 (3.8, 5.5) | 4.26 x 10<sup>-7</sup> |

HP, *Helicobacter pylori*; BMI, body mass index; SBP, systolic blood pressure; DBP, diastolic blood pressure; TC, total cholesterol; TG, triglycerides; HDL, high-density lipoprotein cholesterol; AST (GOT), aspartate aminotransferase (glutamic oxaloacetic transaminase); ALT (GPT), alanine aminotransferase (glutamic pyruvic transaminase). γ-GTP, gamma-glutamyl transpeptidase; Cr, creatinine; UA, uric acid; <sup>a</sup>HP<sup>-</sup> (n=3,154); <sup>b</sup>HP<sup>+</sup> (n=1,824), HP<sup>-</sup> (n=3,153).

**S2 Table.** Continuous variables for participants after matching

| Variable (unit) | HP<sup>+</sup> (n=1,688) | HP<sup>-</sup> (n=1,688) | P value |
| --- | --- | --- | --- |
| Age (yr) | 57.9 (48.6, 64.1) | 57.8 (48.8, 63.9) | 0.866 |
| Waist (cm) | 80.5 (74.6, 87) | 80 (73.9, 86) | 0.0592 |
| SBP (mmHg) | 117 (105.5, 132.5) | 116.5 (105.5, 131) | 0.664 |
| DBP (mmHg) | 71.5 (64, 81.5) | 72 (64.5, 81) | 0.862 |
| TC (mg/dL)<sup>a</sup> | 210 (188, 235) | 211 (188, 235) | 0.983 |
| TG (mg/dL)<sup>a</sup> | 81 (57, 117) | 79 (59, 114) | 0.907 |
| HDL (mg/dL)<sup>a</sup> | 62 (52, 73) | 65 (54, 76) | 0.0000233 |
| AST (GOT) (IU/L)<sup>a</sup> | 20 (17, 24) | 20 (17, 24) | 0.652 |
| ALT (GPT) (IU/L)<sup>a</sup> | 16 (12, 21.5) | 16 (12, 21) | 0.883 |
| γ-GTP (IU/L)<sup>a</sup> | 20 (14, 32) | 20 (14, 31) | 0.997 |
| Cr (mg/dL)<sup>a</sup> | 0.6 (0.6, 0.7) | 0.6 (0.6, 0.7) | 0.836 |
| UA (mg/dL)<sup>a</sup> | 4.7 (4, 5.6) | 4.6 (3.9, 5.6) | 0.144 |

HP, *Helicobacter pylori*; BMI, body mass index; SBP, systolic blood pressure; DBP, diastolic blood pressure; TC, total cholesterol; TG, triglycerides; HDL, high-density lipoprotein cholesterol; AST (GOT), aspartate aminotransferase (glutamic oxaloacetic transaminase); ALT (GPT), alanine aminotransferase (glutamic pyruvic transaminase). γ-GTP, gamma-glutamyl transpeptidase; Cr, creatinine; UA, uric acid. <sup>a</sup>HP<sup>+</sup> (n=1,687), HP<sup>-</sup> (n=1,686).

**S3 Table.** Cancer incidence of patients who are HP^+^ and HP^-^.

|  |  | HP<sup>+</sup> (n=1,688) |  | HP<sup>-</sup> (n=1,688) |  |  |
| --- | --- | --- | --- | --- | --- | --- |
| Variable | Value | n | Frequency | n | Frequency | *P value |
| All cancer | No | 1,583 | 0.938 | 1,621 | 0.96 | 0.00367 |
|  | Yes | 105 | 0.062 | 67 | 0.04 |  |
| Gastric cancer | No | 1,668 | 0.988 | 1,683 | 0.997 | 0.00395 |
|  | Yes | 20 | 0.012 | 5 | 0.003 |  |
| Non-gastric cancer | No | 1,601 | 0.948 | 1,626 | 0.963 | 0.044 |
|  | Yes | 87 | 0.052 | 62 | 0.037 |  |
| Uterine | No | 1,679 | 0.995 | 1,683 | 0.997 | 0.0423 |
|  | Yes | 9 | 0.005 | 5 | 0.003 |  |
| Lung | No | 1,682 | 0.996 | 1,680 | 0.995 | 0.790 |
|  | Yes | 6 | 0.004 | 8 | 0.005 |  |
| Prostate | No | 1,678 | 0.994 | 1,681 | 0.996 | 0.628 |
|  | Yes | 10 | 0.006 | 7 | 0.004 |  |
| Colon | No | 1,672 | 0.991 | 1,685 | 0.996 | 0.092 |
|  | Yes | 16 | 0.009 | 3 | 0.004 |  |
| Breast | No | 1,670 | 0.989 | 1677 | 0.993 | 0.263 |
|  | Yes | 18 | 0.011 | 11 | 0.007 |  |
| **Death | No | 1,661 | 0.984 | 1,661 | 0.984 | 1.00 |
|  | Yes | 27 | 0.016 | 27 | 0.016 |  |

Freq, frequency; \*Fisher's exact test; \*\*all-cause death is not a part of cancer incidence but presented as relevant information.

**S4 Table.** Multivariate Cox regression models for all-cancer incidence (*n*=3,375).

| Variable | HR | 95%CI Lower | 95%CI Upper | P value |
| --- | --- | --- | --- | --- |
| Age (yr) | 1.07 | 1.05 | 1.09 | $3.35 \times 10^{-11}$ |
| Sex | 0.88 | 0.60 | 1.30 | 0.52 |
| Drinking | 1.24 | 0.88 | 1.74 | 0.216 |
| Smoking | 1.97 | 1.35 | 2.87 | 0.000422 |
| HP | 1.59 | 1.17 | 2.16 | 0.00297 |

HR, hazard ratio; HP, *Helicobacter pylori*.

**S5 Table.** Multivariate Cox regression models for gastric cancer incidence (*n*=3,375).

| Variable | HR | 95%CI Lower | 95%CI Upper | P value |
| --- | --- | --- | --- | --- |
| Age | 1.11 | 1.04 | 1.19 | 0.000948 |
| Sex | 0.36 | 0.13 | 1.02 | 0.0542 |
| Drinking | 0.76 | 0.32 | 1.83 | 0.542 |
| Smoking | 1.40 | 0.50 | 3.90 | 0.519 |
| HP | 3.93 | 1.47 | 10.48 | 0.00631 |

HR, hazard ratio; HP, *Helicobacter pylori*.

**S6 Table.** Multivariate Cox regression models for non-gastric cancer incidence (*n*=3,375).

| Variable | HR | 95%CI Lower | 95%CI Upper | P value |
| --- | --- | --- | --- | --- |
| Age | 1.06 | 1.04 | 1.09 | 5.02 x 10<sup>-09</sup> |
| Sex | 0.99 | 0.66 | 1.50 | 0.975 |
| Drinking | 1.31 | 0.91 | 1.89 | 0.147 |
| Smoking | 2.09 | 1.40 | 3.13 | 0.0003 |
| HP | 1.42 | 1.02 | 1.97 | 0.0356 |

HR, hazard ratio; HP, *Helicobacter pylori*.

**S1 Fig.**
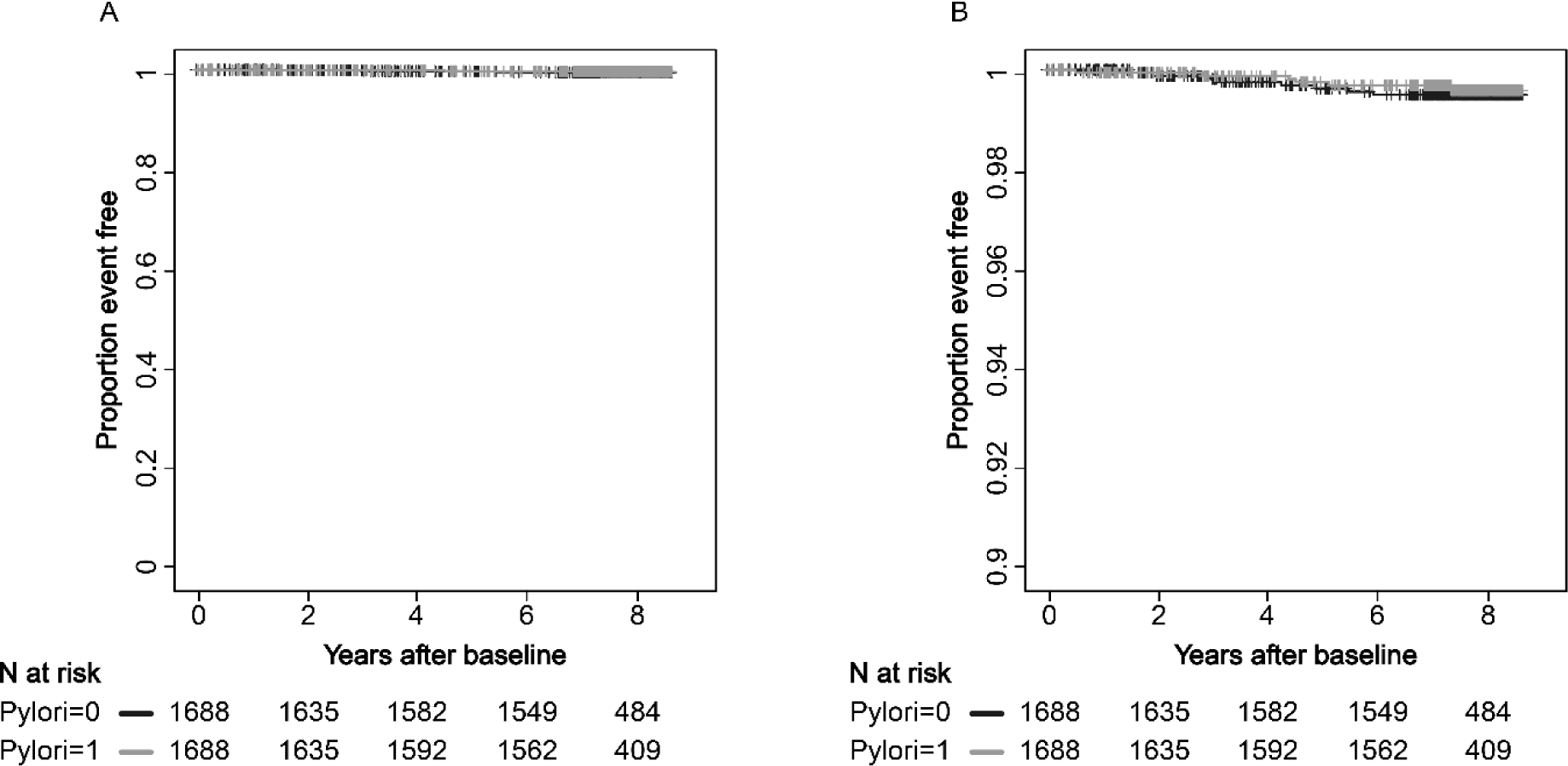
Lung cancer incidence. Kaplan-Meier estimation of lung cancer incidence stratified by anti-HP antibody status. A, axis range 0.00-1.00; B, axis range 0.90-1.00.

**S2 Fig.**
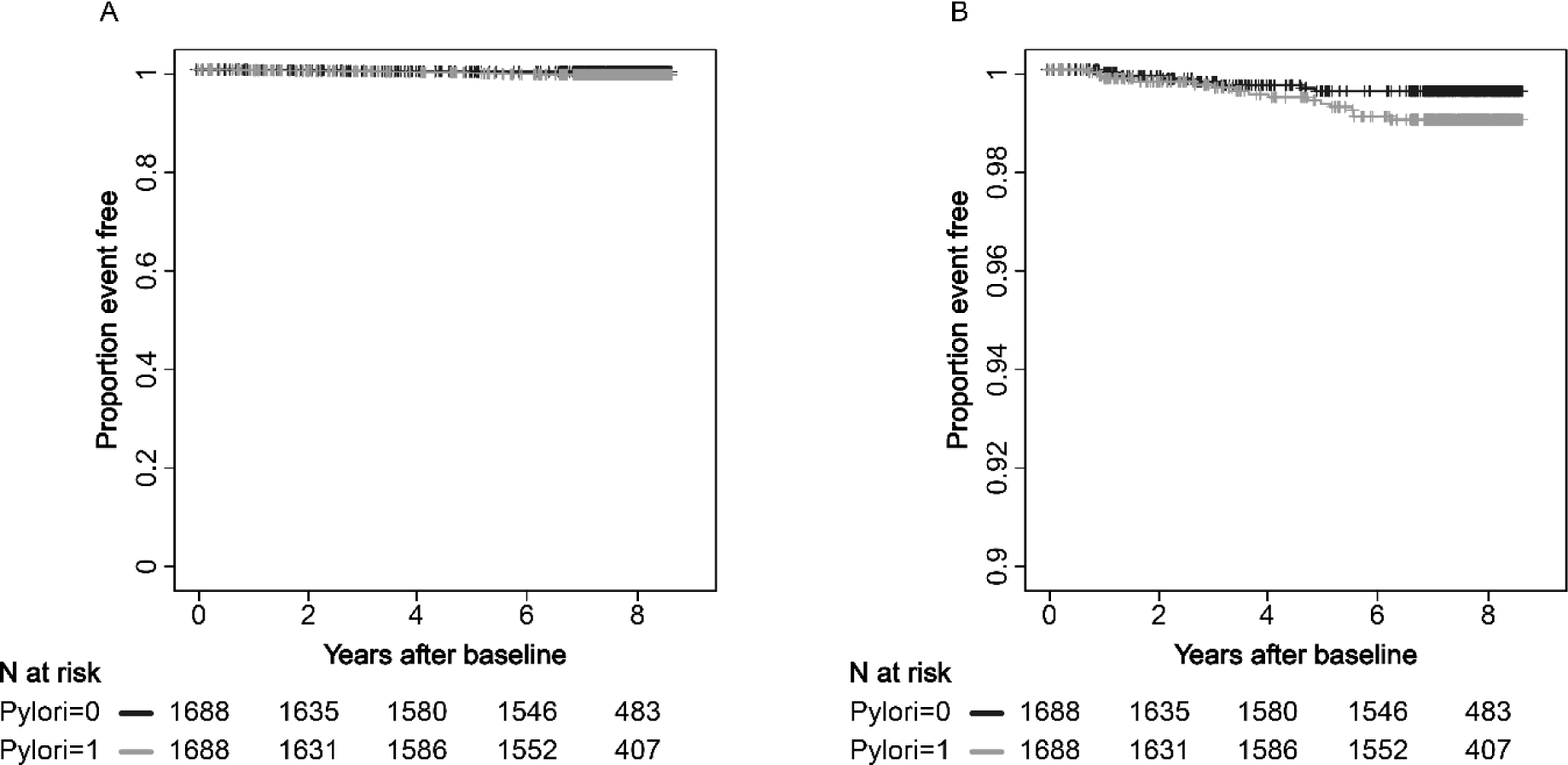
Colorectal cancer incidence. Kaplan-Meier estimation of colorectal cancer incidence stratified by anti-HP antibody status. A, axis range 0.00-1.00; B, axis range 0.90-1.00.

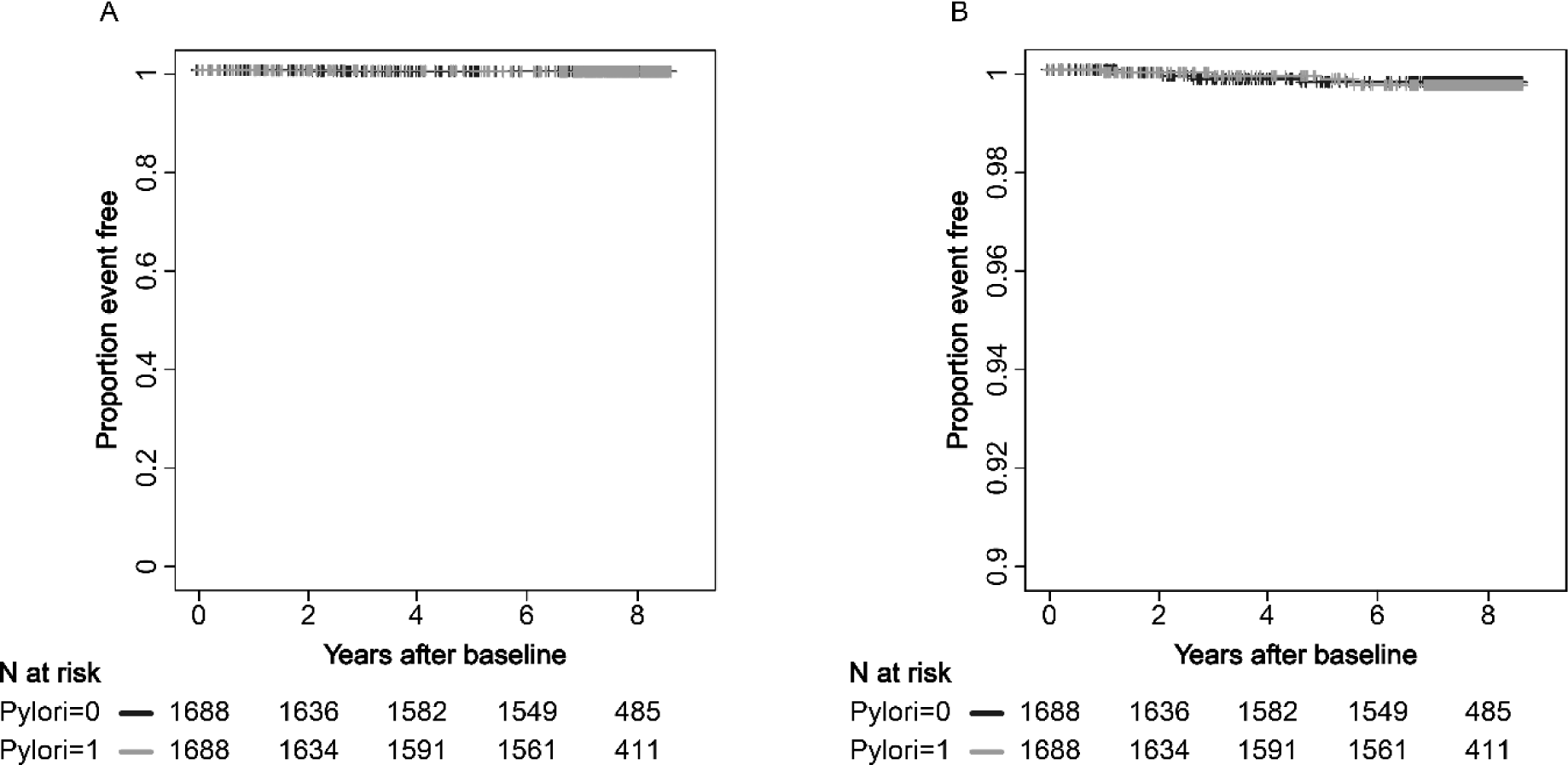
Rectal cancer incidence.

**S4 Fig.**
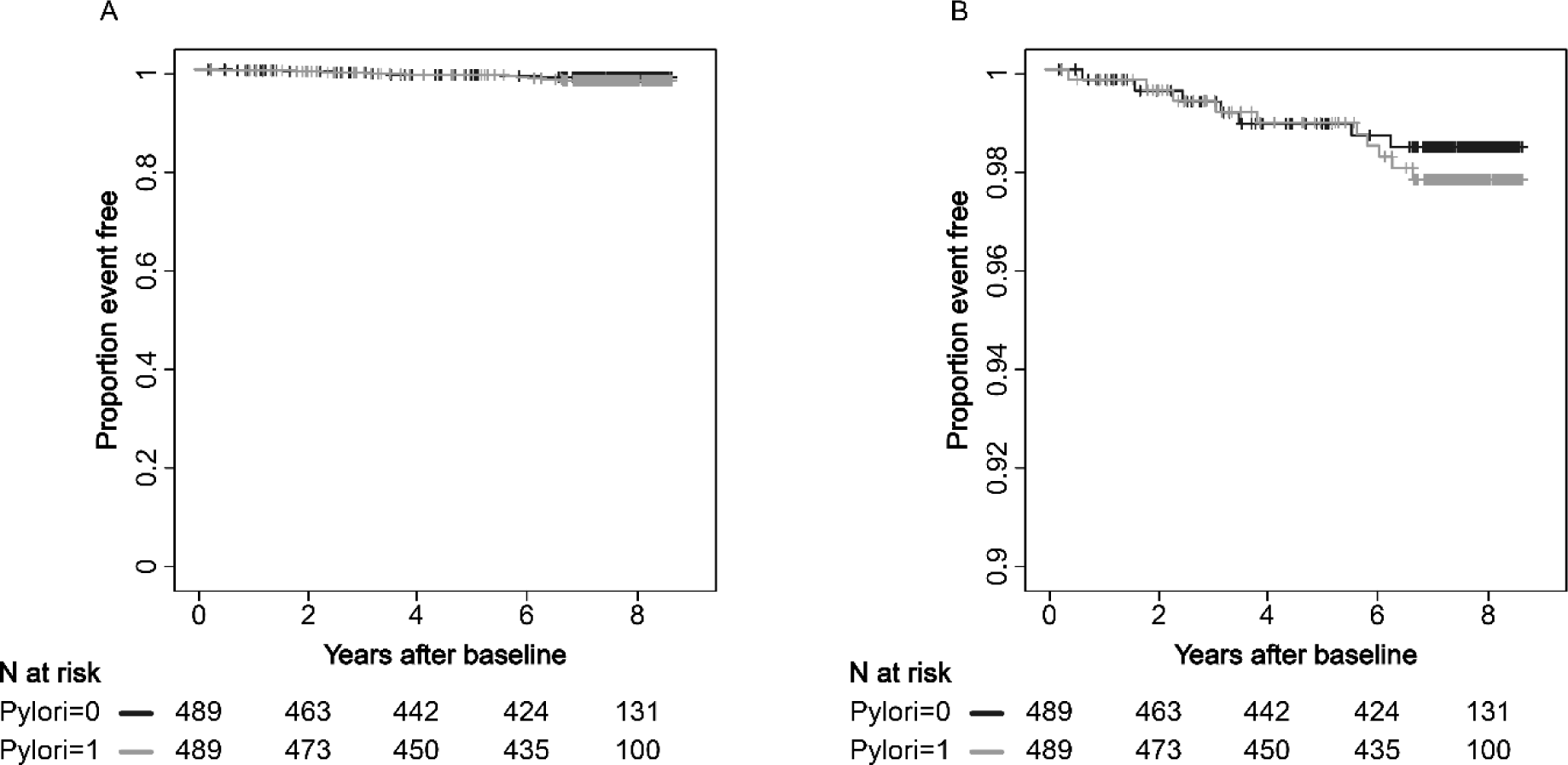
Prostate cancer incidence. Kaplan-Meier estimation of prostate cancer incidence stratified by anti-HP antibody status. A, axis range 0.00-1.00; B, axis range 0.90-1.00.

**S5 Fig.**
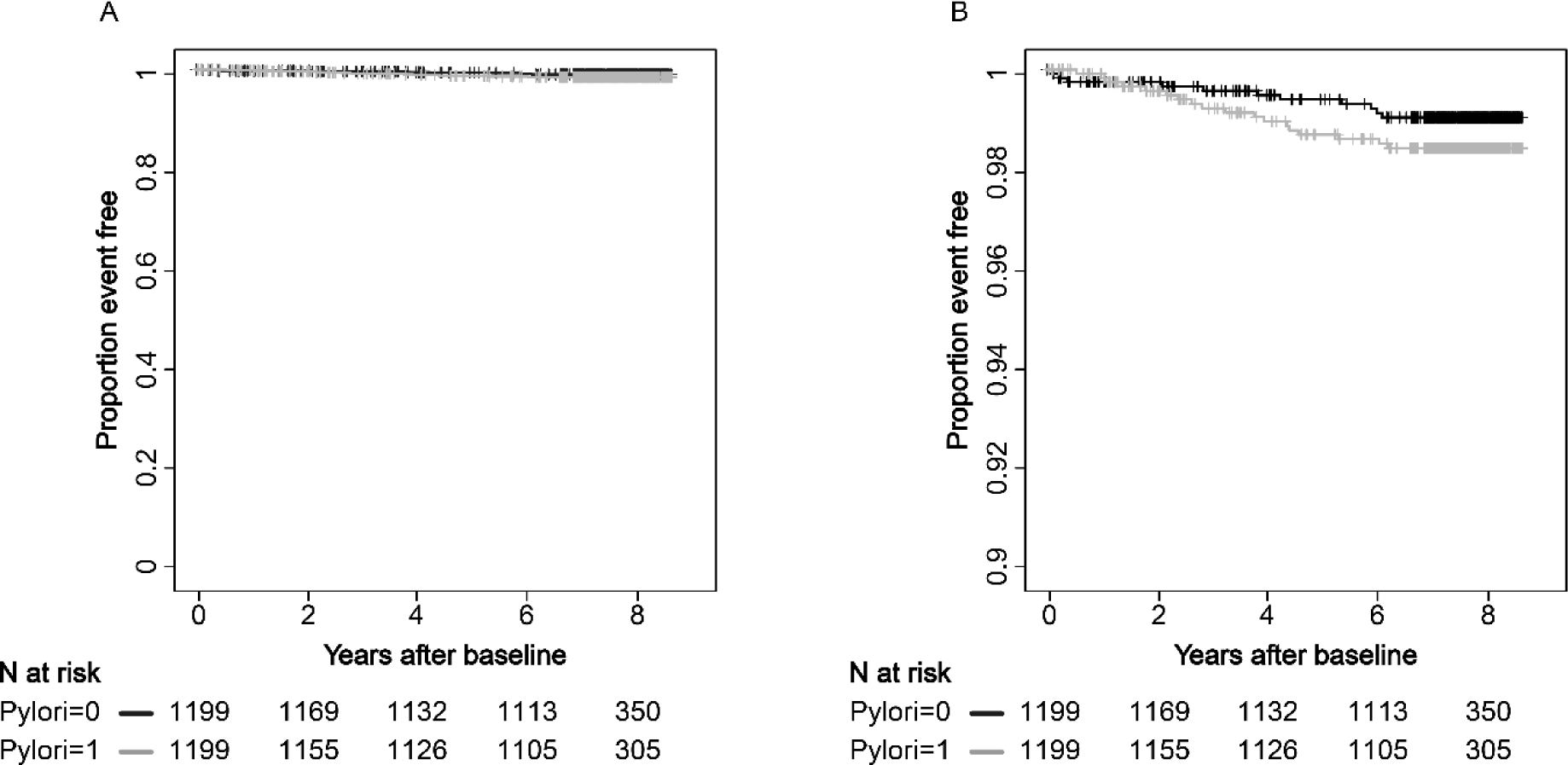
Breast cancer incidence. Kaplan-Meier estimation of lung cancer incidence stratified by anti-HP antibody status. A, axis range 0.00-1.00; B, axis range 0.90-1.00.

## Notes

### Competing Interest Statement

The authors have declared no competing interest.

### Funding Statement

This study was funded by Grants-in-Aid for Scientific Research for Priority Areas of Cancer (No. 17015018); Grants-in-Aid for Innovative Areas (No. 221S0001); and the Japan Society for the Promotion of Science (JSPS) KAKENHI Grant (No. 19K09130 and No. 16H06277 [CoBiA]) from the Japanese Ministry of Education, Culture, Sports, Science and Technology (MEXT). The funders had no role in study design, data collection and analysis, decision to publish, or preparation of the manuscript.

### Author Declarations

Both Nagoya University Graduate School of Medicine IRB (2008-0618-2) and Iwate Medical University School of Medicine IRB (MH2018-019) gave approval for this work.

